# Dyslipidemia as the Predictive Factor for the Severity of Diabetic Retinopathy Among Type 2 Diabetic Patients at KCMC Hospital from 2023 to 2024

**DOI:** 10.1101/2025.07.02.25330746

**Authors:** Muhidini Huud Swalehe, William Makupa, Andrew Makupa

## Abstract

**Background:** Diabetes mellitus (DM) is a global health concern, leading to complications like diabetic retinopathy (DR), a leading cause of blindness. Dyslipidemia, marked by abnormal lipid levels, is considered a risk factor for DR due to its role in endothelial dysfunction and inflammation. However, studies on the relationship between lipid levels and DR severity have yielded conflicting results, making this a debated topic.

**Study Aim:** This study aimed to assess dyslipidemia as a predictor for DR severity in type 2 diabetic patients attending Kilimanjaro Christian Medical Centre (KCMC) from 2023 to 2024.

**Methodology:** A hospital-based analytical cross-sectional study was conducted with 296 diabetic outpatients at KCMC. Participants underwent fundoscopy and were evaluated for DR severity, then blood samples were collected for biochemical analysis. Data were cleaned and analyzed using STATA version 17.

**Results:** Among the patients, 31.4% (93/296) had no DR, while 68.6% (203/296) had DR. Abnormally high levels of serum cholesterol (48.6%), triglycerides (43.6%), LDL (36.1%), and low HDL (38.9%) were identified among study participants. Elevated serum triglycerides and LDL were significantly associated with DR severity (p < 0.05), while serum cholesterol was not significant after adjustment (p = 0.068). Progression from NPDR to PDR was linked to diabetes duration over 5 years and high serum triglycerides (p ≤ 0.001), but HDL levels were not significantly associated with DR. Abnormal BMI was a significant predictor of diabetic retinopathy (p = 0.002), while urine protein and HbA1c levels were not significant after adjustment.

**Conclusion:** The study found a strong link between dyslipidemia and the severity of DR. Even after adjustments LDL, and triglycerides were significantly associated with DR severity. Diabetes duration and Body mass index were also significant factors. The study emphasizes the importance of early intervention, lifestyle changes, enhanced screening programs, and patient education to manage diabetes and its complications and reduce DR incidence and progression.

## Introduction

Diabetes is a persistent, metabolic disorder defined by high levels of blood sugar which leads to long-term severe harm to the heart, blood vessels, eyes, kidneys and nerves. There are two primary forms of Diabetic mellitus, specifically type 1 and type 2. When the body ceases to produce adequate insulin or develops insulin insensitivity, type 2 diabetes occurs and when the pancreas produces insufficient or no insulin naturally, Type 1 diabetes occurs (WHO). The prevalence of type 2 diabetes is increasing gradually due to factors such as increased obesity, reduced physical activity, and sedentary lifestyles (1).

Globally, diabetes mellitus is a significant healthcare issue, with over 360 million individuals estimated to be impacted by the condition by 2030 (2). According to the International Diabetes Federation, the number of diagnosed adults with DM in Africa will rise from 12.1 million in 2010 to 23.9 million in 2030 (3).

Diabetic retinopathy is one of the primary microvascular complications of diabetes and is considered a significant cause of permanent blindness worldwide (4) and among individuals globally with Diabetes Mellitus, 34.6% will develop DR, and 10.2% will progress to vision-threatening DR by 2030 (2). It has been divided into two groups, namely NPDR (Non proliferative diabetic retinopathy) and PDR (proliferative diabetic retinopathy). NPDR is identified by the presence of microaneurysms, cotton-wool spots, venous beading, intraretinal microvascular abnormalities, retinal hemorrhage, and hard exudates in the fundus view. On the other hand, PDR is characterized by the development of neovascularization on the retina surface, which leads to complications such as retinal and vitreous hemorrhages, fibrosis, and tractional retinal detachment (5).

Various factors that increase the risk of its advancement have been identified. The Diabetes Control and Complications Trial revealed that while 11% of DR risks can be attributed to glycaemic exposure, the remaining 89% may be linked to other potential factors (6). The primary risk factors identified were prolonged diabetic duration, hyperglycemia, and hypertension. Additionally, dyslipidemia has also been suggested as a contributing risk factor for diabetic retinopathy (4).

Dyslipidemia refers to an atypical lipid profile marked by elevated levels of serum total cholesterol (TC), triglycerides (TG), low-density lipoprotein cholesterol (LDL-C), and decreased levels of high-density lipoprotein cholesterol (HDL-C) (7), (8) or the presence of one or multiple abnormal levels of lipids in the bloodstream (9).

Globally, dyslipidemia affects approximately 70% to 97% of individuals with diabetes, largely driven by unhealthy diets, obesity, and sedentary lifestyles. Reported prevalence rates are notably high 88.9% in Thailand, 90% in Jordan, 86.1% in Kenya (10) and around 83% in Tanzania (11). Abnormal lipid levels significantly elevate the risk of vascular complications, including diabetic retinopathy, by triggering endothelial dysfunction, oxidative stress through lipid peroxidation, and chronic inflammation mechanisms that collectively accelerate retinal damage (4,7)

Nevertheless, the significance of dyslipidaemia as a self-reliant risk element in the development of DR is debatable. Several investigations have demonstrated a substantial correlation between serum lipids and DR, whereas others have not discovered any notable association. Consequently, the accounts concerning the link between serum dyslipidaemia and DR remain unclear.

Considering the discrepancies in findings across global research and the limited information available in domestic literature, the objective of this study is to investigate the association between serum lipids and diabetic retinopathy in East Africa, and Africa as whole.

## Methodology

A hospital based cross-sectional study aimed to evaluate dyslipidemia as a predictor for DR severity in type 2 diabetic patients attending Kilimanjaro Christian Medical Centre (KCMC) from 2023 to 2024.

The study enrolled a total of 296 patients with Type 2 diabetes mellitus who were attending the Outpatient clinic at Kilimanjaro Christian Medical Centre (KCMC), using a consecutive non-probability sampling method. Patients were excluded if they had retinal vascular conditions such as retinal vein occlusion, were on lipid-lowering medications, were pregnant, had chronic kidney disease, hemolytic disorders, a history of vitreoretinal surgery, media opacities obstructing fundus visualization, or a prior diagnosis of glaucoma.

Diabetic retinopathy was assessed as the dependent variable, while the independent variables included triglyceride levels, total serum cholesterol, high-density lipoprotein (HDL), low-density lipoprotein (LDL), duration of diabetes, sociodemographic characteristics, blood pressure, HbA1c, presence of proteinuria, serum creatinine, and body mass index (BMI).

Data were collected using a structured form divided into five sections: informed consent, demographic details, physical examination, diabetic retinopathy grading, macular edema assessment, and laboratory investigations. Demographic data included patient identification number, age, gender, residence, diabetes duration, treatment type, visual acuity, and intraocular pressure. Laboratory evaluations covered serum cholesterol, triglycerides, HDL, LDL, HbA1c, serum creatinine, and urinalysis.

Ethical approval was obtained from the Kilimanjaro Christian Medical University College Research and Ethics Review Committee (Ref: PG 89/2023), with institutional clearance granted via the Ophthalmology Department at KCMC. All participants were informed about the study’s purpose, and written consent was obtained. Participant confidentiality and data protection were maintained throughout.

After obtaining informed consent, visual acuity (VA) and intraocular pressure (IOP) were measured using a Snellen chart and Goldmann applanation tonometer, respectively. Blood pressure was assessed using a digital sphygmomanometer. Comprehensive fundus examination was performed using indirect ophthalmoscopy with a 90D Volk lens on a Haag-Streit slit lamp, following pupillary dilation with 0.5% tropicamide and 5% phenylephrine. Based on retinal findings and the Diabetic Retinopathy Disease Severity Scale, patients were categorized into: no diabetic retinopathy (NDR), non-proliferative diabetic retinopathy (NPDR), and proliferative diabetic retinopathy (PDR). NDR included eyes without visible fundus abnormalities. NPDR was defined by features such as microaneurysms, dot and blot hemorrhages, IRMA, cotton wool spots, hard exudates, and venous beading. PDR was characterized by neovascularization, vitreous hemorrhage, or proliferative membranes. There after laboratory investigations were conducted, including serum lipid profiling (total cholesterol, triglycerides, HDL, and LDL) using the COBAS INTEGRA 400 PLUS analyzer. A lipid profile was considered abnormal if HDL was <1.15 mmol/L, LDL >3.37 mmol/L, triglycerides >2.26 mmol/L, or total cholesterol >5.2 mmol/L. Additional tests included serum creatinine, glycated hemoglobin (HbA1c), and urinalysis.

Data were analyzed using STATA version 17 (Stata Corp LLC, College Station, TX, USA). Prior to analysis, data cleaning procedures including encoding, labeling, recoding, and variable definition were performed to ensure consistency. Categorical variables were presented as frequencies and percentages, while numerical data were summarized using means with standard deviations or medians with interquartile ranges. The Chi-square (χ²) test was applied to assess differences in DR severity, with a p-value <0.05 considered statistically significant. Ordinal logistic regression was employed to identify factors associated with DR severity. Univariate models were first used to obtain crude odds ratios. Variables with p-values <0.05 and those of clinical relevance were included in multivariable models to compute adjusted odds ratios. Associations with p-values <0.05 in the multivariable analysis were considered statistically significant.

## Results

A total of 296 participants were enrolled, with a mean age of 62 years; 44.6% were male and 55.4% female. The majority (97.3%) had been living with diabetes for more than five years. Severe visual impairment was observed in 34.8% of right eyes and 33.6% of left eyes. Elevated systolic blood pressure (≥140 mmHg) was noted in 61.1% of participants, and 35.1% had diastolic pressure ≥90 mmHg. Additionally, 37.8% were overweight and 12.8% as obese (**Table 1**).

**Table 1:** Social demographic and clinical characteristics of the study participants (N=296)

| **Variable** | **Frequency** | **Percentage** |
| --- | --- | --- |
| **Age** |  |  |
| *Mean (SD)* | 62.4 ($\pm 8.77$) | |
| **Sex** |  |  |
| Male | 132 | 44.6 |
| Female | 164 | **55.4** |
| **Residence** |  |  |
| Urban | 258 | **87.2** |
| Rural | 38 | 12.8 |
| **Duration of DM** |  |  |
| $\leq 5$ Years | 8 | 2.7 |
| $> 5$ Years | 288 | **97.3** |
| *Mean (SD)* | 11 ($\pm 3.53$) | |
| **Type of medicine use** |  |  |
| Oral | 293 | **99** |
| Injectable | 3 | 1 |
| **Visual acuity of Right Eye** |  |  |
| Mild visual impairment | 145 | **49** |
| Moderate visual impairment | 48 | 16.2 |
| Severe visual impairment | 103 | 34.8 |
| **Visual acuity of Left eye** |  |  |
| Mild visual impairment | 148 | **50.2** |
| Moderate visual impairment | 48 | 16.3 |
| Severe visual impairment | 99 | 33.6 |
| **Intraocular pressure of Right eye** |  |  |
| Normal | 288 | **97.3** |
| High | 8 | 2.7 |
| **Intraocular pressure of Left eye** |  |  |
| Normal | 284 | **95.9** |
| High | 12 | 4.1 |
| **Systolic blood pressure** |  |  |
| $< 140$ | 115 | 38.9 |
| $\geq 140$ | 181 | **61.1** |
| **Diastolic blood pressure** |  |  |
| $< 90$ | 192 | 64.9 |
| $\geq 90$ | 104 | **35.1** |
| **BMI** |  |  |
| Underweight | 4 | 1.4 |
| Normal | 142 | 48 |
| Overweight | 112 | **37.8** |
| Obese | 38 | 12.8 |

In terms of laboratory results, abnormal lipid profiles were prevalent with 48.6% of participants had elevated total cholesterol, 43.6% had high triglycerides, and 36.1% exhibited increased LDL levels, while 38.9% had low HDL levels. Furthermore, glycemic control was poor, with 99% of participants showing HbA1c levels exceeding 5.81% (**Table 2**).

**Table 2:**
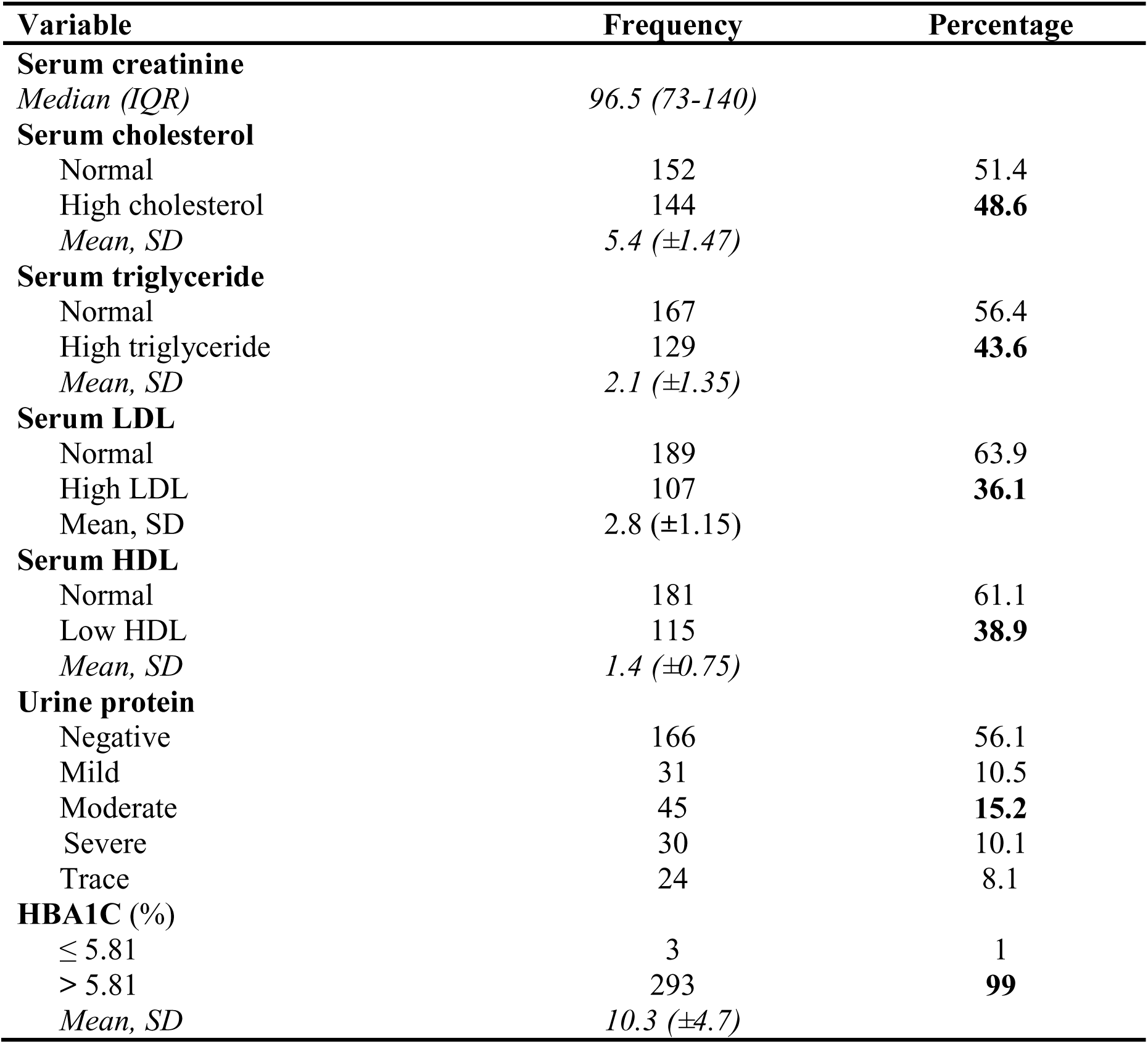
Participants laboratory findings (N=296)

The prevalence of diabetic retinopathy was observed to be 68.6%, with non-proliferative diabetic retinopathy (NPDR) and proliferative diabetic retinopathy (PDR) accounting for 34.5% (102/296) and 34.1% (101/296), respectively. In comparison, 31.4% (93/296) of individuals had no diabetic retinopathy (NDR) **(Figure 1**).

**Figure 1:**
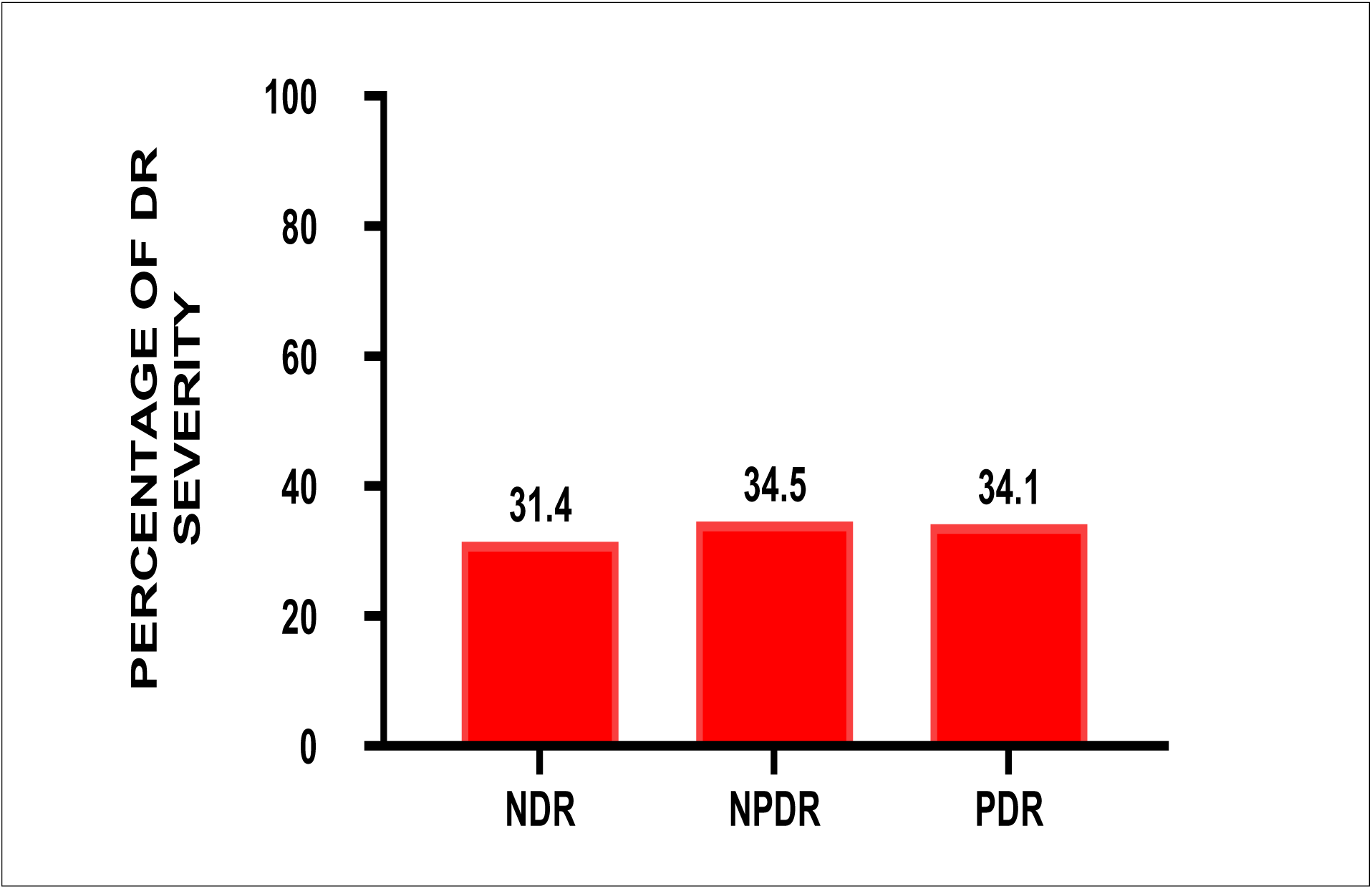
Proportion of Diabetic retinopathy among Diabetes patients (N=296)

In assessing factors associated with the onset of diabetic retinopathy, elevated serum triglyceride levels remained a significant predictor even after adjustment, with a 3.5-fold increased risk of developing NPDR and PDR (AOR = 3.49; 95% CI: 2.12–5.75; p < 0.001). Similarly, elevated LDL levels were linked to a 2.3-fold higher risk (AOR = 2.30; 95% CI: 1.36–3.88; p = 0.021). Total cholesterol and HDL levels were not significantly associated after adjustment. However, both BMI and duration of diabetes remained strong independent predictors (**Table 3**).

**Table 3:** Factors associated with severity of diabetic retinopathy among type 2 diabetic patients (N-296).

| Variable | COR (95%CI) | P-value | AOR (95%CI) | P-value |
| --- | --- | --- | --- | --- |
| **Sex** |  |  |  |  |
| Male | Ref |  |  |  |
| Female | 0.99 (0.65-1.51) | 0.988 |  |  |
| Age | 0.98 (0.97-1.01) | 0.398 |  |  |
| **Duration of DM** | 1.20 (1.12-1.29) | **< 0.001** | 1.23 (1.13-1.33) | **< 0.001** |
| **BMI** | 1.5 (1.13-2.04) | **0.005** | 1.47 (1.06-2.03) | **0.002** |
| **Serum creatinine** | 0.99 (0.997-1.00) | 0.215 |  |  |
| **Serum cholesterol** |  |  |  |  |
| Normal | Ref |  | Ref |  |
| High cholesterol | 2.55 (1.65-3.99) | **< 0.001** | 1.60 (0.97-2.62) | 0.068 |
| **Serum triglycerides** |  |  |  |  |
| Normal | Ref |  | Ref |  |
| High triglyceride | 5.12 (3.23-8.12) | **< 0.001** | 3.49 (2.12-5.75) | **< 0.001** |
| **Serum HDL** |  |  |  |  |
| Low HDL | 1.07 (0.70-1.64) | 0.747 | 0.99 (0.62-1.59) | 0.975 |
| Normal | Ref |  | Ref |  |
| **Serum LDL** |  |  |  |  |
| Normal | Ref |  | Ref |  |
| High LDL | 3.66 (2.32-5.77) | **< 0.001** | 2.30 (1.36-3.88) | **0.021** |
| **Urinalysis protein** |  |  |  |  |
| Normal | Ref |  |  |  |
| Abnormal | 1.48 (0.96 - 2.31) | 0.149 |  |  |
| **HBA1C** | 1.14 (1.04-1.24) | **0.004** | 1.06 (0.97-1.16) | 0.202 |

Among patients already diagnosed with diabetic retinopathy, elevated serum triglycerides were significantly associated with progression from NPDR to PDR, increasing the risk by 65% (APR = 1.65; 95% CI: 1.24–2.21; p = 0.001) while the duration of diabetes also remained as a significant factor in disease progression (**Table 4**).

**Table 4:** Predictive factors for the progression from non-proliferative (NPDR) to proliferative diabetic retinopathy (PDR), (N-203).

| Variable | CPR (95%CI) | P-value | APR (95%CI) | P-value |
| --- | --- | --- | --- | --- |
| **Sex** |  |  |  |  |
| Male | Ref |  |  |  |
| Female | 1.04 (0.79-1.37) | 0.778 |  |  |
| Age | 0.99 (0.97-1.01) | 0.260 |  |  |
| **Duration of diabetic mellitus** | 1.11 (1.08-1.16) | **<0.001** | 1.12(1.08-1.16) | **<0.001** |
| **BMI** | 1.02 (0.98-1.05) | 0.297 |  |  |
| **Serum creatinine** | 1.00 (0.99-1.001) | 0.246 |  |  |
| **Serum cholesterol** |  |  |  |  |
| < 5.2 | Ref |  | Ref |  |
| ≥ 5.2 | 1.02 (0.77-1.35) | 0.887 | 0.97 (0.74-1.28) | 0.872 |
| **Serum triglycerides** |  |  |  |  |
| < 2.26 | Ref |  | Ref |  |
| ≥ 2.26 | 1.58 (1.16-2.15) | **0.003** | 1.65 (1.24-2.21) | **0.001** |
| **Serum HDL** |  |  |  |  |
| ≤ 1.15 | 0.94 (0.70-1.25) | 0.668 | 0.96 (0.74-1.25) | 0.769 |
| > 1.15 | Ref |  | Ref |  |
| **Serum LDL** |  |  |  |  |
| < 3.37 | Ref |  | Ref |  |
| ≥ 3.37 | 1.10 (0.91-1.58) | 0.206 | 1.09 (0.83-1.43) | 0.528 |
| **Urinalysis protein** |  |  |  |  |
| Normal | Ref |  |  |  |
| Abnormal | 1.20 (0.91-1.58) | 0.189 |  |  |
| **HBA1C** | 1.05 (0.99-1.11) | 0.080 |  |  |

## Discussion

In this study, a considerable number of participants had abnormal lipid profiles: 48.6% had elevated cholesterol, 43.6% had high triglycerides, 36.1% had increased LDL, and 38.9% had reduced HDL levels. These findings may reflect poor dietary practices, physical inactivity, and limited healthcare access. In comparison, a study in Bangladesh reported even higher rates 52.3% for cholesterol, 57.6% for triglycerides, 50% for LDL, and 74.2% for low HDL) (12). Conversely, Rwanda recorded a much lower dyslipidemia prevalence of 19.2% (13).These disparities may be partly influenced by genetic variations, including genes such as APOE, PCSK9, and LPL, which regulate lipid metabolism and contribute to dyslipidemia risk.

Diabetic retinopathy was highly prevalent in our study, affecting 68.6% of participants, with 31.4% showing no signs of retinopathy (NDR). This higher prevalence could be attributed to factors like longer diabetes duration and poor glycemic control both established risk factors for advanced retinopathy. Additionally, the use of advanced diagnostic tools (e.g., OCT), referral bias to tertiary care centers, specialized services for severe cases, and socioeconomic barriers to timely healthcare may have influenced detection rates. These findings align with (4) in India, where NDR was 39.7% and diabetic retinopathy 60.3%. Both studies share similar single-center, hospital-based settings, with patients having a mean diabetes duration of over 10 years. However, our results differ from those of (14) in Bangalore and (15) in Southern Ethiopia, where NDR proportions were higher, likely due to the younger age of their participants.

A significant association was observed between dyslipidemia and increased severity of diabetic retinopathy. Elevated serum cholesterol, triglycerides, and LDL levels were linked to higher odds of developing NPDR and PDR. This may be attributed to mechanisms such as endothelial dysfunction, lipid peroxidation, induced oxidative stress, and inflammation, all of which contribute to retinopathy progression. These results align with previous studies, including (2) in India, who reported significantly higher lipid levels among patients with diabetic retinopathy. However, our findings contrast with those of (7) who found no significant association, possibly due to their smaller sample size.

In examining predictors for progression from NPDR to PDR, a strong independent association was found with both diabetes duration and lipid profile abnormalities. Each additional year of diabetes was linked to a 12% increased risk of developing PDR, likely reflecting the cumulative effects of chronic hyperglycemia on retinal microvasculature, consistent with findings by (16) in Pakistan. Among lipid markers, elevated serum triglycerides showed the strongest association, conferring a 65% higher risk of PDR. Similar observations were reported by (17) in Saudi Arabia, where participants had comparable mean ages.

Interestingly, this study found no significant association between HDL levels and the severity of diabetic retinopathy. This may be attributed to population-based variations in HDL subclasses (HDL2 and HDL3), as also reported by (18) in Guntur, India. Conversely, (16) in Pakistan observed a significant association between reduced HDL and increased retinopathy severity. These inconsistencies could be influenced by geographic differences and the younger mean age of participants in their study compared to ours.

This study also found a significant association between higher BMI and increased severity of diabetic retinopathy. This is likely due to the link between elevated BMI and insulin resistance, which impairs glycemic control and contributes to chronic hyperglycemia a known driver of retinopathy progression. Our findings are consistent with those of (19) in Croatia, where similar associations were observed, possibly due to a high prevalence of hypertension in both cohorts. These results underscore the potential role of metabolic syndrome in retinal vascular damage and highlight the need for further investigation in this area.

A key limitation of this study is its hospital-based design, which may limit the generalizability of findings to the broader population of individuals with type 2 diabetes. Patients attending tertiary care centers are more likely to present with advanced disease or complications, potentially leading to an overestimation of the prevalence and severity of diabetic retinopathy and dyslipidemia. To address this in future research, population-based studies involving community-level screening and broader sampling strategies are recommended to provide a more representative picture of disease patterns and associated risk factors.

## Conclusion

The study revealed a strong connection between dyslipidemia and the severity of diabetic retinopathy. Even after adjustments, LDL, and triglycerides were significantly associated with the severity of diabetic retinopathy. Additionally, the duration of diabetes and BMI remained significant factors. These findings underscore the importance of early intervention, lifestyle modification, and metabolic control in reducing the risk of vision-threatening complications. Furthermore, the study highlights the need for enhanced screening programs and patient education to support timely detection and management of diabetic retinopathy.

## Recommendations

Future research should prioritize large-scale, well-designed studies such as case-control designs to better establish the causal relationship between dyslipidemia and diabetic retinopathy progression. Meanwhile, clinical practice should incorporate routine lipid screening, patient education, and lifestyle interventions into diabetes care. Strengthened interdisciplinary collaboration and supportive national policies are also essential to ensure early detection, effective lipid management, and the prevention of vision-threatening diabetic complications.

## Data Availability

All data produced in the present study are available upon reasonable request to the authors

## Acknowledgement

I thank Dr. William Makupa and Dr. Andrew Makupa for their invaluable mentorship and guidance.

I sincerely appreciate Justus Rwiza, Kalvin Rwegoshora for his support in formal analysis, data analysis and biostatistics.

Grateful to CBM for financial support and to my family for their constant encouragement.

## Authors contribution

Conceptualization: Muhidini Huud Swalehe.

Formal analysis: Muhidini Huud Swalehe, Justus Rwiza, Kevin Rwegoshola.

Funding acquisition: Christian Blind Mission (CBM)

Investigation: Muhidini Huud Swalehe

Methodology: Muhidini Huud Swalehe

Project administration: Muhidini Huud Swalehe

Writing original draft: Muhidini Huud Swalehe.

Review and editing: Muhidini Huud Swalehe, William Makupa, Andrew Makupa

